## Supplementary material for "Hemodynamic Differences Between Women and Men with Elevated Blood Pressure in China: A Non-Invasive Assessment of 45,082 Adults Using Impedance Cardiography": S1 Fig

**Short title: Hemodynamic Sex Differences in Elevated Blood Pressure**

**S1 Fig. Median Cardiac Output, Cardiac Index, Systemic Vascular Resistance, and Systemic Vascular Resistance Index by Age Among Women and Men with Systolic Blood Pressure ≥140 mmHg or Diastolic Blood Pressure ≥90 mmHg.**

**S2 Fig. Cardiac Output and Cardiac Index Density Plots Overlap Between Women and Men with Systolic Blood Pressure ≥140 mmHg or Diastolic Blood Pressure ≥90 mmHg, by Age Category.**

**S3 Fig. Systemic Vascular Resistance and Systemic Vascular Resistance Index Density Plots Overlap Between Women and Men with Systolic Blood Pressure ≥140 mmHg or Diastolic Blood Pressure ≥90 mmHg, by Age Category.**

**S1 Table. Sex Differences in Clinical and Hemodynamic Variables by Age Group Among Adults with Systolic Blood Pressure ≥140 mmHg or Diastolic Blood Pressure ≥90 mmHg.**

**S2 Table. Sex Differences in Clinical and Hemodynamic Variables by Nearest Neighbor Propensity Score Matched Subgroups.**

**S3 Table. Unadjusted and Sequentially-Adjusted Association of Female Sex with Cardiac Output, Cardiac Index, Systemic Vascular Resistance, and Systemic Vascular Resistance Index, Overall and by Age Categories Among Adults Systolic Blood Pressure ≥140 mmHg or Diastolic Blood Pressure ≥90 mmHg, by Age Category.**

**S1 Figure.** Median Cardiac Output, Cardiac Index, Systemic Vascular Resistance, and Systemic Vascular Resistance Index by Age Among Women and Men with Systolic Blood Pressure ≥140 mmHg or Diastolic Blood Pressure ≥90 mmHg.

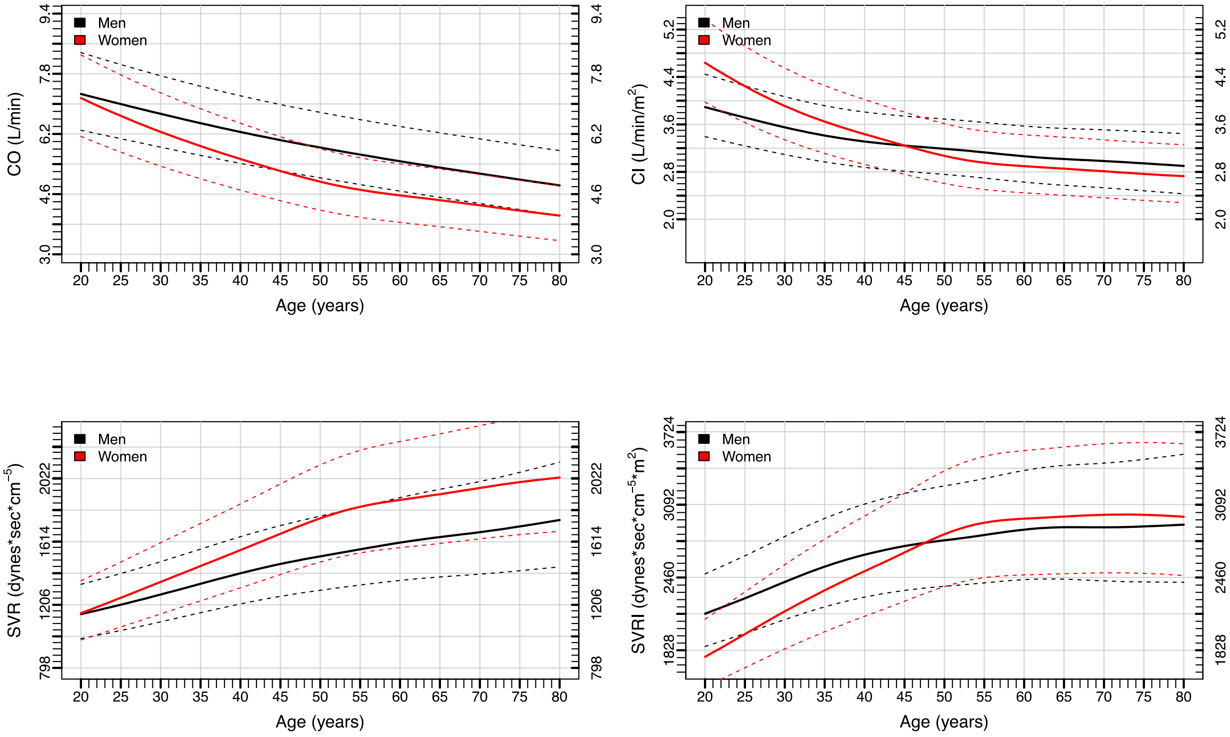


Solid lines represent the median. Dashed lines represent the $25^{th}$ and $75^{th}$ percentile. Abbreviations: CO, cardiac output; CI, cardiac index; SVR, systemic vascular resistance; SVRI, systemic vascular resistance index.

**S2 Figure.** Cardiac Output and Cardiac Index Density Plots Overlap Between Women and Men with Systolic Blood Pressure ≥140 mmHg or Diastolic Blood Pressure ≥90 mmHg, by Age Category.

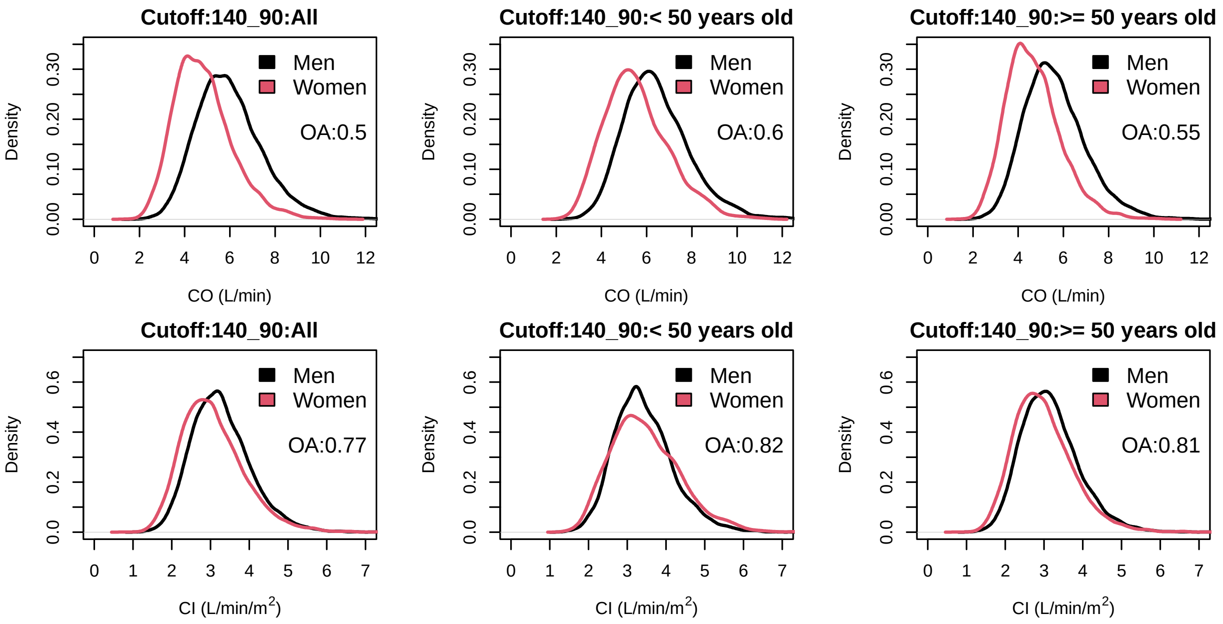


CO, cardiac output; CI, cardiac index; OA, overlapping area.

**S3 Figure.** Systemic Vascular Resistance and Systemic Vascular Resistance Index Density Plots Overlap Between Women and Men with Systolic Blood Pressure ≥140 mmHg or Diastolic Blood Pressure ≥90 mmHg, by Age Category.

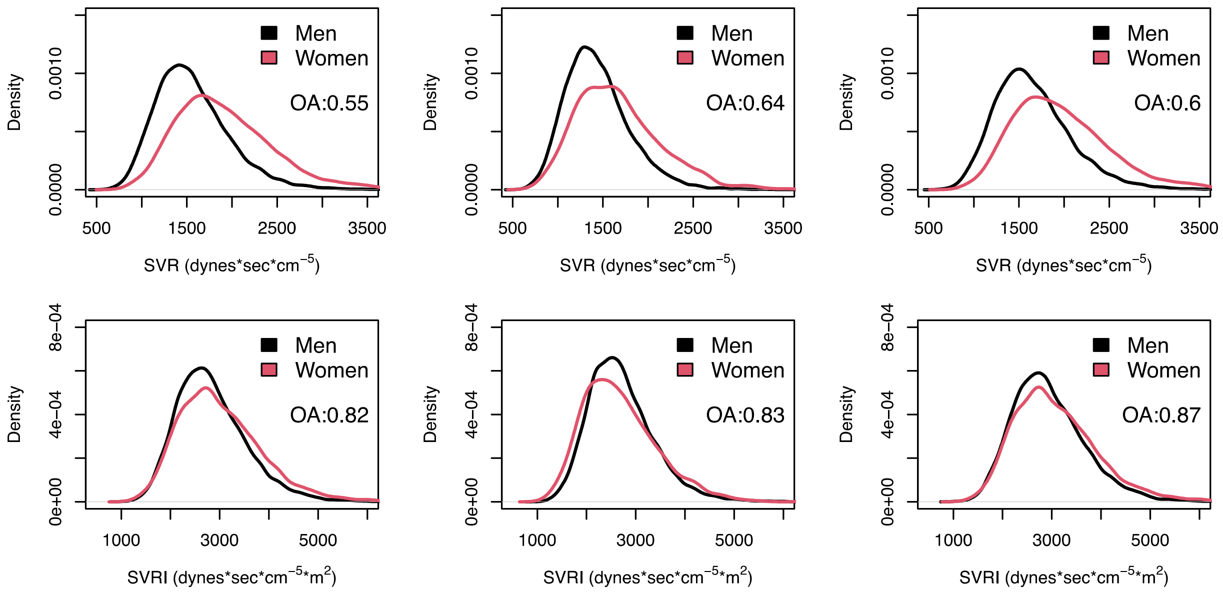


SVR, systemic vascular resistance; SVRI, systemic vascular resistance index; OA, overlapping area.

**S1 Table .** Sex Differences in Clinical and Hemodynamic Variables By Age Group Among Adults with Systolic Blood Pressure ≥140 mmHg or Diastolic Blood Pressure ≥90 mmHg.

| | **All** | | | **< 50 years old** | | | **≥ 50 years old** | | |
|---|---|---|---|---|---|---|---|---|---|
| | Women N=7593 | Men N=12711 | P value | Women N = 1246 | Men N = 5439 | P value | Women N = 6347 | Men N = 7272 | P value |
| Age (years), mean (SD) | 58.13 (10.02) | 51.27 (12.72) | < 0.001 | 42.1 (6.85) | 39.22 (7.11) | < 0.001 | 61.27 (7.12) | 60.29 (7.43) | < 0.001 |
| BMI (kg/m2), mean (SD) | 25.02 (3.51) | 25.89 (3.29) | < 0.001 | 24.63 (3.84) | 26.49 (3.51) | < 0.001 | 25.1 (3.44) | 25.44 (3.05) | < 0.001 |
| Obese (BMI ≥27.5 kg/m2), n(%) | 1623 (21.37%) | 3560 (28.01%) | < 0.001 | 248 (19.9%) | 1884 (34.64%) | < 0.001 | 1375 (21.66%) | 1676 (23.05%) | 0.06 |
| Region, n (%) | | | < 0.001 | | | < 0.001 | | | <0.001 |
| East | 2776 (36.56%) | 5834 (45.9%) | | 542 (43.5%) | 2653 (48.78%) | | 2234 (35.2%) | 3181 (43.74%) | |
| North | 1737 (22.88%) | 1869 (14.7%) | | 219 (17.58%) | 669 (12.3%) | | 1518 (23.92%) | 1200 (16.5%) | |
| South | 1360 (17.91%) | 1792 (14.1%) | | 200 (16.05%) | 785 (14.43%) | | 1160 (18.28%) | 1007 (13.85%) | |
| Southwest | 1720 (22.65%) | 3216 (25.3%) | | 285 (22.87%) | 1332 (24.49%) | | 1435 (22.61%) | 1884 (25.91%) | |
| Blood pressure in mmHg, mean (SD) | | | | | | | | | |
| Systolic | 150.75 (13.68) | 147.48 (13.02) | < 0.001 | 144.68 (13.71) | 144.48 (12.18) | 0.60 | 151.94 (13.36) | 149.72 (13.18) | < 0.001 |
| Diastolic | 86.31 (10.17) | 91.43 (9.46) | < 0.001 | 90.5 (8.98) | 92.97 (9.25) | < 0.001 | 85.49 (10.19) | 90.28 (9.44) | < 0.001 |
| Hypertension phenotype, n (%) | | | | | | | | | |
| Predominantly cardiac (High CI with low/normal SVRI) | 923 (12.16%) | 1957 (15.40%) | < 0.001 | 318 (25.52%) | 1063 (19.54%) | < 0.001 | 263 (4.14%) | 894 (12.29%) | < 0.001 |
| Predominantly vascular (Low/normal CI with high SVRI) | 5671 (74.69%) | 8952 (70.43%) | < 0.001 | 732 (58.75%) | 3474 (63.87%) | < 0.001 | 4939 (77.82%) | 5478 (75.33%) | < 0.001 |
| Low/normal CI & SVRI | 981 (12.92%) | 1737 (13.67%) | 0.14 | 190 (15.25%) | 872 (16.03%) | 0.522 | 791 (12.46%) | 865 (11.89%) | 0.33 |
| High CI & SVRI | 18 (0.24%) | 65 (0.51%) | 0.004 | 6 (0.48%) | 30 (0.55%) | 0.928 | 12 (0.19%) | 35 (0.48%) | 0.006 |
| ICG parameters, mean (SD) | | | | | | | | | |
| Heart rate (bpm) | 69.69 (11.47) | 70.49 (11.66) | < 0.001 | 73.27 (11.99) | 72.47 (11.53) | 0.03 | 68.99 (11.23) | 69.01 (11.54) | 0.91 |
| Stroke volume (mL) | 70.01 (17.95) | 86.01 (21.19) | < 0.001 | 76.73 (18.25) | 90.22 (21.48) | < 0.001 | 68.69 (17.59) | 82.86 (20.42) | < 0.001 |
| CO (L/min) | 4.83 (1.29) | 5.98 (1.47) | < 0.001 | 5.57 (1.42) | 6.44 (1.46) | < 0.001 | 4.68 (1.21) | 5.63 (1.37) | < 0.001 |
| CI ($L/min/m^2$) | 3.06 (0.8) | 3.27 (0.76) | < 0.001 | 3.47 (0.87) | 3.42 (0.75) | 0.06 | 2.99 (0.76) | 3.16 (0.75) | < 0.001 |
| SVR ($dynes \cdot sec \cdot cm^{-5}$) | 1917.42 (549.88) | 1564.95 (417.38) | < 0.001 | 1666.98 (468.62) | 1442.02 (363.25) | < 0.001 | 1966.59 (551.29) | 1656.9 (431.37) | < 0.001 |

| | | | | | | | | | |
|---|---|---|---|---|---|---|---|---|---|
| SVRI (dynes·sec·cm$^{-5}$·m$^{2}$) | 3011.81 (850.4) | 2843.28 (726.62) | < 0.001 | 2674.73 (746.86) | 2707.12 (675.2) | 0.14 | 3077.98 (853.85) | 2945.1 (746.81) | < 0.001 |
| SD= Standard Deviation, BMI= Body Mass Index, ICG= Impedance Cardiography, SVR= Systemic Vascular Resistance, SVRI= Systemic Vascular Resistance Index, CO= Cardiac Output, CI= Cardiac Index. | | | | | | | | | |

**S2 Table.** Sex Differences in Clinical and Hemodynamic Variables by Nearest Neighbor Propensity Score Matched Subgroups.

| | All<br>1:1 Matching | | | < 50 years old<br>1:1 Matching | | | ≥ 50 years old<br>1:1 Matching | | |
|---|---|---|---|---|---|---|---|---|---|
| | Women<br>N=15,888 | Men<br>N= 15,888 | P value | Women<br>N = 4,384 | Men<br>N = 4,384 | P value | Women<br>N = 11,504 | Men<br>N = 11,504 | P value |
| Age, years mean (SD) | 54.46 (11.83) | 53.45 (12.72) | <0.001 | 39.3 (8.07) | 39.27 (7.5) | 0.84 | 60.24 (6.9) | 59.68 (7.24) | <0.001 |
| BMI (kg/m2), mean (SD) | 24.4 (3.49) | 24.53 (2.95) | <0.001 | 23.49 (3.72) | 23.65 (3.3) | 0.04 | 24.75 (3.33) | 24.97 (2.94) | <0.001 |
| Obese (BMI ≥27.5 kg/m2), n(%) | 2733 (17.2%) | 2284 (14.38%) | <0.001 | 585 (13.34%) | 483 (11.02%) | <0.001 | 2148 (18.67%) | 2086 (18.13%) | 0.30 |
| Region, n (%) | | | <0.001 | | | 0.26 | | | <0.001 |
| East | 5965 (48.74%) | 6273 (51.26%) | | 1956 (50.46%) | 1920 (49.54%) | | 4009 (46.58%) | 4597 (53.42%) | |
| North | 3665 (51.84%) | 3405 (48.16%) | | 779 (47.94%) | 846 (52.06%) | | 2886 (55.98%) | 2269 (44.02%) | |
| South | 2737 (50.39%) | 2695 (49.61%) | | 686 (49.67%) | 695 (50.33%) | | 2051 (52.32%) | 1869 (47.68%) | |
| South West | 3521 (50.04%) | 3515 (49.96%) | | 963 (51.06%) | 923 (48.94%) | | 2558 (48.02%) | 2769 (51.98%) | |
| Blood pressure in mmHg, mean (SD) | | | | | | | | | |
| Systolic | 139.02 (15.71) | 138 (13.98) | <0.001 | 131.21 (13.19) | 131.52 (11.06) | 0.04 | 142 (15.57) | 140.36 (14.57) | <0.001 |
| Diastolic | 82.64 (9.00) | 83.62 (8.39) | <0.001 | 83.32 (7.85) | 83.37 (8.05) | 0.23 | 82.37 (9.38) | 84.61 (8.32) | <0.001 |
| ICG parameters, mean (SD) | | | | | | | | | |
| CO (L/min) | 5.01 (1.35) | 5.87 (1.43) | <0.001 | 5.75 (1.41) | 6.39 (1.42) | <0.001 | 4.73 (1.21) | 5.63 (1.35) | <0.001 |
| CI (L/min/m2) | 3.19 (0.84) | 3.31 (0.78) | <0.001 | 3.64 (0.88) | 3.59 (0.77) | 0.013 | 3.02 (0.76) | 3.18 (0.74) | <0.001 |
| SVR<br>(dynes·sec·cm-5) | 1743.94 (523.42) | 1475.6 (406.58) | <0.001 | 1470.91 (411.34) | 1306.85 (315.19) | <0.001 | 1847.99 (524.09) | 1552.76 (410.64) | <0.001 |
| SVRI<br>(dynes·sec·cm-5·m2) | 2734.1 (809.92) | 2606.44 (691.7) | <0.001 | 2325.95 (658.03) | 2322.25 (556.02) | 0.78 | 2889.64 (808.27) | 2741.28 (702.93) | <0.001 |

SD= Standard Deviation, BMI= Body Mass Index, ICG= Impedance Cardiography, SVR= Systemic Vascular Resistance, SVRI= Systemic Vascular Resistance Index, CO= Cardiac Output, CI= Cardiac Index.
Propensity Score Generation Model: Gender ~ Age + BMI + SBP + DBP + Region

**S3 Table.** Unadjusted and Sequentially-Adjusted Association of Female Sex with Cardiac Output, Cardiac Index, Systemic Vascular Resistance, and Systemic Vascular Resistance Index, Overall and by Age Categories Among Adults Systolic Blood Pressure ≥140 mmHg or Diastolic Blood Pressure ≥90 mmHg, by Age Category.

| **Hemodynamic Variable** | **Female Sex β Coefficient (95% CI)** | | |
|---|---|---|---|
| | Unadjusted Model | Adjusted Model 1* | Adjusted Model 2** |
| **Cardiac Output,** (L/min) | | | |
| Overall | -1.15 (-1.19, -1.11) | -0.87 (-0.91, -0.84) | -0.87 (-0.91, -0.83) |
| <50 years old | -0.87 (-0.96, -0.78) | -0.71 (-0.8, -0.63) | -0.67 (-0.76, -0.58) |
| ≥50 years old | -0.95 (-1, -0.91) | -0.91 (-0.96, -0.87) | -0.91 (-0.96, -0.87) |
| **Cardiac Index,** (L/min/$m^2$) | | | |
| Overall | -0.21 (-0.23, -0.19) | -0.09 (-0.11, -0.07) | -0.13 (-0.16, -0.11) |
| <50 years old | 0.05 (0, 0.09)[a] | 0.13 (0.08, 0.17) | 0.02 (-0.03, 0.06)[b] |
| ≥50 years old | -0.17 (-0.2, -0.15) | -0.15 (-0.18, -0.13) | -0.17 (-0.2, -0.15) |
| **Systemic Vascular Resistance,** (dynes·sec·$cm^{-5}$) | | | |
| Overall | 352.47 (339.07, 365.87) | 273.29 (259.87, 286.72) | 275.51 (262.02, 289) |
| <50 years old | 224.95 (201.24, 248.66) | 181.64 (158.55, 204.72) | 183.61 (159.99, 207.23) |
| ≥50 years old | 309.69 (293.16, 326.22) | 298.18 (281.67, 314.69) | 299.21 (282.67, 315.74) |
| **Systemic Vascular Resistance Index,** (dynes·sec·$cm^{-5}$·$m^2$) | | | |
| Overall | 168.52 (146.48, 190.56) | 67.26 (44.85, 89.68) | 112.69 (90.99, 134.39) |
| <50 years old | -32.39 (-74.82, 10.04)[c] | -108.78 (-150.29, -67.26) | 1.48 (-39.14, 42.09)[d] |
| ≥50 years old | 132.85 (105.97, 159.74) | 114.16 (87.29, 141.03) | 136.87 (110.84, 162.89) |

* Model 1 was adjusted for age and region
** Model 2 was adjusted for age, region, and body mass index

[a]P value=0.06
[b]P value=0.51
[c]P value=0.13
[d]P value=0.94
All other P values <0.001
